# Factors associated with health decision-making autonomy on own healthcare among Tanzanian women: A 2022-2023 demographic health survey study

**DOI:** 10.1101/2024.04.01.24305186

**Authors:** Tausi Haruna, Henry Duah, Rebecca Lee

## Abstract

**Background:** Women’s health decision-making autonomy is fundamental for the health and well-being of women and their children. Like most parts of Africa, women’s status in Tanzania remains contentious, with an estimated 19% prevalence of health decision-making autonomy in 2015. Given that women’s health decision-making autonomy is an ongoing process affected by technological and economic growth and social and cultural changes, understanding the sociodemographic correlates of women’s autonomy is imperative.

**Objective:** To examine the factors associated with health decision-making autonomy on their own health among Tanzanian women aged 15-49.

**Methods:** A non-experimental cross-sectional study using secondary data from the current Tanzania Demographic and Health Survey and Malaria Indicator Survey (TDHS-MIS) 2022-2023. The R statistical programming language was used to run the analysis. Chi-square and Ordinal Logistic Regression were fitted to identify the sociodemographic characteristics associated with women’s health decision-making autonomy on their own health. The odds ratio with its 95% confidence interval was used to determine the significance level at p-value <0.05. All estimates were adjusted for sample design (sample weight, strata, and sampling units)

**Results:** A total of 9,249 women were included in the analysis. Only 1,908 (21%) had complete autonomy, 4,933 (53%) had joint autonomy, and 2,408 (26%) had no autonomy. Women’s age, education level, household wealth index, and living in the South West Highlands zone were factors independently associated with higher odds of complete autonomy in their own healthcare. Rural residence was associated with decreased odds of complete autonomy.

**Conclusion:** These results show that health decision-making autonomy among Tanzanian women remains very low. Efforts to empower women through better education and means to improve their economic status are needed to increase complete health decision-making autonomy on their health.

**Recommendation:** Accelerated and concerted efforts to increase health decision-making autonomy among married women will eventually improve their health and well-being and that of society.

**Future implication:** The findings can serve as a basis for exploratory qualitative research to further understand the process of health decision-making autonomy among Tanzanian women.

## Introduction

Women’s health decision-making autonomy is fundamental for the health and well-being of women and their children. It facilitates access to knowledge, power, and socio-cultural resources within the family and community. Women’s autonomy in healthcare decision-making is one of the indicators of women’s health outcomes and empowerment [1,2]. The Sustainable Development Goals (SDGs) set a target to reduce maternal mortality to less than 70 deaths per 100,000 live births in 2023. Without paying attention to women’s health decision-making autonomy, efforts to achieve this goal and improve women’s health outcomes may be compromised [3].

Women’s autonomy is a multifaceted concept comprising cultural, social structures, and economic dimensions [4]. Because of its multidimensional nature, there is no single acceptable definition of women’s autonomy. Studies on women’s autonomy defined it according to context, situation, and purpose [5,6]. Dyson and Moore, 1983 referred to women’s autonomy as *the technical, social, and psychological ability to obtain information and use it as the basis for making decisions about one’s private concerns and those of one’s intimates* [7]. Basu defined women’s autonomy as the capacity and freedom to act independently, for instance, the ability to go places, such as health facilities or markets, or to make decisions regarding contraceptive use or household purchases alone and without asking anyone’s permission [8]. Mason defined it as women’s ability to make and execute independent decisions pertaining to personal matters of importance to their lives and families [9]. With regard to the healthcare context, women’s autonomy in this study is defined as the ability of women to make independent decisions on their own healthcare [10].

Existing studies in low and middle-income countries (LMICs) show a close correlation between women’s socio-demographic characteristics and health decision-making autonomy [11–14]. A study among Nigerian women examining the perceived health decision-making autonomy about their own healthcare found that geographical region, rural/urban residence, age, education, religion, wealth index, and occupation independently associated with women’s health decision-making autonomy [15]. Nigatu et al., 2014 and Singh et al.,2015, reported that women with a higher wealth index are more likely to have choices regarding their own healthcare compared to their counterparts [5,16]. In Tanzania, a study by Masawe et al., 2019 found that women with secondary and post-secondary education, older and employed, tend to be more autonomous [14]. Similar findings were reported in Nepal, where women with increased education, employment, and a higher number of living children were associated with increased autonomy in decision-making [13]. Few studies in Tanzania have addressed the dynamics associated with decision-making autonomy, including the various country’s zones.

Like most parts of Africa, women’s status in Tanzania remains contentious, with an estimated 19% prevalence of health decision-making autonomy in 2015 [14]. This signifies that women have low autonomy in health decision-making. Decisions on maternal healthcare are often made with their husbands/partners or someone else, which negatively influences maternal and child healthcare utilization [17]. Government, non-government, and international efforts that promote women’s autonomy in health decision-making have been employed [18]. The Tanzanian government has been striving to implement policies that promote equality and women’s rights and encourage the participation of women in decision-making [19]. The government also enforces strategies that promote access to education for girls and women and enhance women’s participation in higher education and economic empowerment by focusing on programs that seek to improve women’s financial independence [18].

Considering all these efforts and given that women’s health decision-making autonomy is an ongoing process affected by technological and economic growth and social and cultural changes, understanding the sociodemographic correlates of women’s autonomy is imperative. Therefore, this study examined the relationship between health decision-making autonomy and sociodemographic characteristics among Tanzanian women of reproductive age 15-49. This relationship becomes particularly important as various programs in low-income countries, including Tanzania, seek to improve women’s health outcomes by increasing women’s autonomy in health decision-making. The study addressed the following questions: 1) To what extent and whether socio-demographic characteristics (age, education, residence, wealth index, and zones of country) are independently associated with health decision-making autonomy? The study hypotheses were: 1) There is a significant association between sociodemographic factors and health decision-making autonomy. 2) There are regional/zone differences in Tanzanian women’s decision-making autonomy about their own healthcare.

## Method

### Design

This was a secondary data analysis of the Tanzania Demographic and Health Survey and Malaria Indicator Survey (TDHS-MIS) 2022. Data from the TDHS-MIS 2022 individual recode documentation were specifically used for this study. The TDHS-MIS is a nationwide survey conducted every five years. The original TDHS-MIS 2022 was undertaken by the Tanzanian National Bureau of Statistics (NBS) and the Office of the Chief Government Statistician Zanzibar (OCGS) in collaboration with affiliate governmental agencies, including the Ministries of Health (MoH) of Tanzania, both in the Mainland and Zanzibar, and the Tanzania Food and Nutrition Centre (TFNC) amongst other agencies. Technical assistance for the survey was provided by ICF international. The 2022 TDHS-MIS was undertaken using a multistage sampling design. The initial phase involved the selection of clusters (primary sampling units), which comprised enumeration areas defined during the 2012 Population and Housing Census. In all, 629 clusters were selected in phase 1, comprising 211 and 418 from urban and rural areas, respectively. The second phase involved systematically selecting households from the selected clusters during phase 1. Averagely, 26 households were selected from each cluster, making a projected total of 16,354 households for the survey. All women in their reproductive age (15-49 years) in selected households were eligible for participation, and those who consented were interviewed on a range of individual and population health issues, including themes on health decision-making autonomy.

### Characteristics of participants

All women of reproductive age from all regions in Tanzania. According to the World Health Organization (WHO), women of reproductive age are all women aged 15-49 years married or in union [3]. Study participants were selected to fit the eligibility criteria under the study: 1) women who were married or living with a partner at the time of the interview, 2) responded to the question, who usually decides on the respondent’s healthcare. A total of 9,249 women met these criteria.

### Study variables and measurements

#### Outcome Variable

The outcome variable for this study was women’s health decision-making autonomy. This variable was measured by using survey questionnaires in the TDHS-MIS data. This was obtained from the variable “decisions on personal health care.” Women were asked, “Who usually decides on the respondent’s healthcare?” Five responses were recorded, “responded alone,” “respondent and partner,” “husband/partner alone,” “someone else,” and “other.” This was recoded at trilevel as “respondent alone =complete autonomy =1”, “respondent and partner =joint autonomy =2”, “partner alone =no autonomy =4”, and “someone else =no autonomy=5”. Respondents who answered “others” were dropped. Therefore, the three levels of autonomy considered in this study were “*No Autonomy,” “Joint Autonomy,”* and “*Complete Autonomy.* For this study, a woman’s autonomy was defined as the capacity of the woman to decide alone on their healthcare [10].

#### Independent Variables

The key explanatory variables for this study were the respondent’s sociodemographic characteristics. The woman’s age, level of education, place of residence, household wealth index, and country’s zones. All explanatory variables were categorical variables. The wealth index in the DHS data was a composite variable measure of a household’s cumulative living standard and relative wealth, calculated using data on the household’s ownership of selected assets, including types of water access and sanitation facilities, televisions and bicycles, and materials for housing construction among others assets. The resulting index was then presented in ordered categories: poorest, poorer, middle, richer, and richest. For statistical analysis, several variables were recoded into categories. The woman’s age was recoded into quinary age groups: 15-19 years, 20-24 years, 25-29 years, 30-34 years, 35-39 years, 40-44 years, and 45-49 years. Women aged 15-19 years were a reference group. The woman’s level of education was recoded as no education =0, primary =1, secondary =2, higher =3, whereas no education was a reference group: place of residence recoded as urban= 1, rural =2; urban was a reference group: household wealth index as poorest =1, poorer =2, middle=3, richer=4, and richest =5; whereas poorest was our reference group: and zones were recoded as Western =1, Northen=2, Central =3, Southern Highlands =4, Southern =5, Southwest Highlands =6, Lake=7, Eastern =8, and Zanzibar =9: the Western zone was the reference group.

### Statistical Analysis

The R statistical programming language was used to perform the analysis. Data were imported into R programming language and cleaned using the appropriate packages in R. Descriptive statistics were calculated using frequencies. A chi-square was used to describe the sociodemographic characteristics of the respondents and their association with women’s health decision-making autonomy on their own health (See Table 1). A multivariable Ordinal Logistic Regression was used to assess the independent association between the explanatory and outcome variables. The variable selection in the regression model was determined by their significance associated with health decision-making autonomy at the bivariate analysis or as reported in the previous studies [14,15]. The Ordinal Logistic Regression was conducted using a single multivariable model. The coefficients were exponentiated to derive adjusted odds ratios (AORs) and corresponding 95% Confidence Intervals (CI) estimates. Statistically significant results were pegged at 0.05 alpha level. All univariate, bivariate, and multivariable analyses accounted for complex survey design.

**Table 1.** Sociodemographic Characteristics of Participants (N=9249)

| Variable | Frequency (n) | Percentage (%) |
| --- | --- | --- |
| **Age -years** |  |  |
| 15-19 | 563 | 6.1 |
| 20-24 | 1,612 | 17 |
| 25-29 | 1,894 | 20 |
| 30-34 | 1,616 | 17 |
| 35-39 | 1,427 | 15 |
| 40-44 | 1,181 | 13 |
| 45-49 | 954 | 10 |
| **Place of residence** |  |  |
| Urban | 2,894 | 31 |
| Rural | 6,355 | 69 |
| **Education** |  |  |
| No education | 1,887 | 20 |
| Primary | 5,395 | 58 |
| Secondary | 1,861 | 20 |
| Higher | 106 | 1.1 |
| **Wealth index** |  |  |
| Poorest | 1715 | 19 |
| Poorer | 1713 | 19 |
| Middle | 1760 | 19 |
| Richer | 1970 | 21 |
| Richest | 2090 | 23 |
| **Zones** |  |  |
| Western | 806 | 8.7 |
| Northern | 1058 | 11 |
| Central | 948 | 10 |
| Southern Highlands | 541 | 5.8 |
| Southern | 453 | 4.9 |
| Southwest Highlands | 862 | 9.3 |
| Lake | 2774 | 30 |
| Eastern | 1519 | 16 |
| Zanzibar | 287 | 3.1 |
| **Health decision making autonomy** |  |  |
| No autonomy | 2408 | 26 |
| Joint autonomy | 4933 | 53 |
| Complete autonomy | 1908 | 21 |

### Ethical approval

The ethical approval for the DHS was the Institutional Review Board (IRB) of ICF International and the National Institute of Medical Research (NIMR), Tanzania. Field agents obtained informed consent from all eligible women prior to enrollment in the survey. The PI was granted permission to use the de-identified secondary data through online registration at the DHS website as https://www.dhsprogram.com/. No additional consent was required from participants in this secondary data analysis.

## Results

### Participants characteristics

A total sample size of 9,249 participants was included in this study. The participants were all married females aged 15-49 years drawn from 31 regions of Tanzania Mainland, and Zanzibar. A large proportion (20%) of women aged 25-29. The majority of women reside in rural areas, 69%. About 58% of women had attained a primary education. Most of the women came from the Lake zone (30%). It was observed that 23% of women were from the richest households. (See Table 1)

### Health decision-making autonomy about own healthcare

Only 21% of women reported being completely autonomous regarding decisions on their own health care, while the majority (53%) of women reported that health decisions about their own healthcare were made jointly, and 26% reported health decisions were made by husbands/partners alone and someone else.

### Factors associated with health decision-making autonomy on own healthcare

Chi-square was conducted to answer whether a relationship exists between sociodemographic characteristics and women’s health decision-making autonomy. Results show that age, education level, place of residence, level of household wealth index, and country’s zones were statistically significantly associated with health decision-making autonomy. See Table 2

**Table 2.** Factors related to health decision-making autonomy on own healthcare (N=9249)

| **Variable** | **No autonomy**<br>**2409 <sup>1</sup>** | **Joint autonomy**<br>**4933 <sup>1</sup>** | **Complete**<br>**autonomy**<br>**1908 <sup>1</sup>** | **p-**<br>**value<sup>2</sup>** |
| --- | --- | --- | --- | --- |
| **Age -years** |  |  |  | **<.001** |
| 15-19 | 227 (9.4) | 269 (5.5) | 68 (3.5) |  |
| 20-24 | 532 (22) | 803 (16) | 277 (15) |  |
| 25-29 | 496 (21) | 1083 (22) | 315 (17) |  |
| 30-34 | 382 (16) | 896 (18) | 337 (18) |  |
| 35-39 | 351 (15) | 718 (15) | 357 (19) |  |
| 40-44 | 220 (9.1) | 633 (13) | 329 (17) |  |
| 45-49 | 199 (8.3) | 530 (11) | 224 (12) |  |
| **Place of residence** |  |  |  | **<.001** |
| Urban | 471 (20) | 1600(32) | 823( 43) |  |
| Rural | 1937 (80) | 3333(68) | 1084 (57) |  |
| **Education** |  |  |  | **<.001** |
| No education | 767 (32) | 784 (16) | 335 (18) |  |
| Primary | 1341 (56) | 2959 (60) | 1095 (57) |  |
| Secondary | 297 (12) | 1121 (23) | 442 (23) |  |
| Higher | 2 (<.1) | 68 (1.4) | 36 (1.9) |  |
| **Wealth index** |  |  |  | **<.001** |
| Poorest | 717 (30) | 740 (15) | 258 (14) |  |
| Poorer | 534(22) | 873 (18) | 307 (16) |  |
| Middle | 452(19) | 1000 (20) | 309 (16) |  |
| Richer | 396(16) | 1139 (23) | 435 (23) |  |
| Richest | 308(13) | 1182 (24) | 599 (31) |  |
| **Zones** |  |  |  | **<.001** |
| Western | 484 (20) | 217 (4.4) | 105 (5.5) |  |
| Northern | 212 (8.8) | 613 (12) | 234 (12) |  |
| Central | 153 (6.4) | 582 (12) | 213 (11) |  |
| Southern Highlands | 50 (2.1) | 423 (8.6) | 68 (3.6) |  |
| Southern | 62 (2.6) | 321 (6.5) | 71 (3.7) |  |
| Southwest Highlands | 84 (3.5) | 575 (12) | 202 (11) |  |
| Lake | 892 (37) | 1318 (27) | 563 (30) |  |
| Eastern | 407 (17) | 739 (15) | 373 (20) |  |
| Zanzibar | 64 (2.7) | 144 (2.9) | 79 (4.1) |  |

Note: <sup>2</sup>=chi-squared test with Rao and Scott's second order of correction; <sup>1</sup>=n (%)

### Multivariable Ordinal Logistic Regression

Three categories were formed depending on how women’s autonomy in health decision-making is constructed in the literature [5,6]. In our analysis, we were interested in modeling the predictors for complete autonomy as a higher level of health decision-making autonomy among women. Therefore, the ordinal logistic regression was conducted to specify the higher hierarchy of women’s health decision-making autonomy, i.e., complete autonomy.

Generally, the odds of having compelete autonomy were greater with increasing age of women. For example, the odds of women having complete health decision-making autonomy was 2.53 times greater for women who were 40-44 years old compared to those 15-19 years old, holding other variables constant. Likewise, the odds of having complete autonomy was greater with increasing levels of formal education relative to no formal educations. For example, the odds of women having complete autonomy was 2.39 times greater for women who had attained higher education than those with no formal education. Similarly, the odds of having complete autonomy was greater with increasing levels of household wealth index. For instance, the results showed that the odds of being completely autonomous increased by 81% among women from the richest households relative to their counterparts from the poorest households. There were zonal variations autonomy status of women in Tanzania. Notably, the odds of having complete autonomy were 5.82 times greater for women from the Southwest Highlands compared to the Western zone. On the contrary, women who resided in rural areas had 38% reduced odds of having complete autonomy compared to their urban counterparts. See Table 3 for more details.

**Table 3.** Multivariate Ordinal Logistic Regression on factors related to women’s health decision-making autonomy on their own healthcare.

| Variable | Unadjusted OR | 95% CI |
| --- | --- | --- |
| **Age -years** |  |  |
| 15-19 | 1 |  |
| 20-24 | 1.28 | [1.04, 1.57] |
| 25-29 | 1.43 | [1.15, 1.77] |
| 30-34 | 1.72 | [1.39, 2.14] |
| 35-39 | 2.06 | [1.62, 2.59] |
| 40-44 | 2.53 | [2.01, 3.18] |
| 45-49 | 2.18 | [1.73, 2.75] |
| **Place of residence** |  |  |
| Urban | 1 |  |
| Rural | 0.62 | [0.49, 0.78] |
| **Education** |  |  |
| No education | 1 |  |
| Primary | 1.38 | [1.17, 1.63] |
| Secondary | 1.72 | [1.38, 2.14] |
| Higher | 2.39 | [1.61, 3.55] |
| **Wealth index** |  |  |
| Poorest | 1 |  |
| Poorer | 1.44 | [1.21, 1.71] |
| Middle | 1.39 | [1.16, 1.67] |
| Richer | 1.63 | [1.34, 1.98] |
| Richest | 1.81 | [1.41, 2.33] |
| **Zones** |  |  |
| Western | 1 |  |
| Northern | 4.00 | [3.03, 5.29] |
| Central | 4.98 | [3.71, 6.67] |
| Southern Highlands | 3.64 | [2.85, 4.67] |
| Southern | 4.13 | [3.11, 5.49] |
| Southwest Highlands | 5.82 | [4.48, 7.58] |
| Lake | 2.66 | [2.08, 3.40] |
| Eastern | 2.35 | [1.77, 3.14] |
| Zanzibar | 3.16 | [2.42, 4.12] |

## Discussion

This study sought to determine whether women’s sociodemographic characteristics correlated with decision-making autonomy on their own health. The results show that women’s age, level of education, household wealth index, and country’s zones independently correlated with health decision-making autonomy on their own health. The odds of being completely autonomous in health decision-making increased with age, formal education level, and household wealth index. The country’s zones were also a determinant factor for women’s complete autonomy in health decision-making. Women living in rural areas had reduced complete autonomy in health decision-making on their own health compared to their counterparts. Furthermore, the findings showed only 21% of Tanzanian women were completely autonomous in health decision-making on own health. It was, however, noteworthy that the majority of the women’s health decision-making was done with husbands/partners (53%) and with someone else (24%).

A study conducted in Tanzania by Masawe et al., (2019) reported a 19% prevalence of health decision-making autonomy among married women [14]. The current study established a 21% prevalence of complete health decision-making autonomy among women. This implied that the women in Tanzania still have limited complete autonomy in making health decisions on their own healthcare. This may be partly attributed to social and cultural norms, religious beliefs, and values influencing how women live and interact with their husbands/partners [20]. It can also be due to the social constructs and gender roles that many developing countries like Tanzania practice that restrict women’s full participation in decisions that affect their health [1,2]. A similar pattern was observed in South Asian countries (Nepal, Bangladesh, and India), where only a small proportion of women (13.4%, 17.6%, and 21.1%, respectively) made health decisions on their healthcare [21].

Women’s age was positively associated with complete autonomy in their health decision-making. Similar findings have been reported in many studies using a national representative sample like ours in LMICs [13–15]. In Nepal, Bangladesh, and India, as women’s age increases, the autonomy in household decisions autonomy increases [21]. The possible reasons associated with this increase include that as women get older, their level of maturity increases, and therefore, they gain more independence in decisions and power [5,13].

This study also found a strong association between women’s level of education and complete autonomy in healthcare decisions. It was observed that as the level of formal education increases, the chances of being completely autonomous in one’s health decisions increased. Higher-educated women were twice as likely to have complete autonomy than those without. This finding is consistent with a study in Nigeria, where higher-educated women were 1.57 times more likely to make decisions alone than those without [15].

The household wealth index affects women’s health decision-making autonomy. Women from richest households were 1.81 times likely to make decisions alone than women from poorest households. This is similar to the studies conducted in Ethiopia and Nigeria [5,15]. In Ghana, it was reported that the wealth index influences maternal healthcare utilization [22,23].

The strongest magnitude of association observed with women’s decision-making autonomy was the country zones. As we hypothesized, country zones were an important determinant for Tanzanian women’s decision-making autonomy, even after controlling for other factors. Tanzania is divided into nine zones (Western, Northern, Central, Southern Highlands, Southern, Southwest Highlands, Lake, Eastern, and Zanzibar). Women living in the Southwest Highlands were five times more likely to decide alone on matters affecting their health compared with the Western zones. This study lays the groundwork for this particular finding. A study conducted in Tanzania by Masawe et al. (2019) did not assess the country’s zone as a determinant for women’s health decision-making autonomy in their own healthcare. Other studies in African countries [15,24–26] have also found that geographical locations were independently associated with women’s autonomy. These findings imply that there are factors associated with geographical locations that are yet to be explicated. A study in India reported that women in the Southern regions of India have more exposure to the outside world, a greater voice in family life, and more freedom of movement than those in the north [8,27]. Osamor and Grady (2018) found that Nigerian women from the South West region were 8.3 times more likely to make their own healthcare decisions than women from the North West region [15]. Very little is known about the influence of zones and women’s health decisions in Tanzania. But family life in Tanzania, like most parts of African countries, is guided by descriptive norms and belief systems that derive from culture and ethnicity. Cultural norms, social norms, and beliefs are central components of regional variations that may partly explain the independent influence of region on women’s autonomy.

Women who resided in rural areas were significantly less likely to make their own health decisions than those from urban areas. These findings were consistent with others conducted in LMICs [5,13–15]. The role of place of residence in decision-making has now been documented beyond doubt in affecting the health of individuals living there. In Tanzania, about (66%) of the population live in rural areas. The geographic isolation of rural populations and the challenges they face in meeting basic social services, economic opportunities, and healthcare services are profound.

### Strengths and limitations

This study has several strengths and weaknesses. One key strength is the representative nature of the data among the Tanzanian women population. To the author’s best knowledge, this is the first study in Tanzania using nationally representative data to establish the findings on the country’s zone as the strongest determinant of women’s health decision-making autonomy in their own healthcare.

The study had the following limitations. Because this was a cross-sectional study, a causal relationship between sociodemographic characteristics (explanatory variables) and health decision-making autonomy (outcome variable) can not be established. Therefore, the conclusion of this paper was based on the association relationship between the explanatory and outcome variables. Since this study was a secondary data analysis, the responses were subjected to recall biases. Also, the nature of the survey questions (closed-ended) has limited the ability to capture the context and nuance of the responses. For example, in this study, we interpreted health decisions made by a woman alone as the higher hierarchy representing women’s autonomy, which could also indicate a lack of support from the husband/partner. To assert this interpretation appropriately, follow-up qualitative prompts are needed to clarify this ambiguity.

## Conclusion

The results show that health decision-making autonomy on healthcare among Tanzanian women remains very low. More than half of married women in Tanzania reported having no capacity to decide on matters related to their own healthcare. Factors associated with women’s health decision-making autonomy include; women’s age, level of education, place of residence, household wealth index, and the country’s zones. Interventions and policies targeting modifiable factors such as education and income-generating activities that increase women’s decision-making autonomy will improve their health outcomes and that of the family and community. Further qualitative research are needed to explores how women interact and navigate various components of health decisions (maternal health, clinic visits, blood transfusion, etc).

## Recommendations

Accelerated and concerted efforts to increase health decision-making autonomy among married women will eventually improve their health and well-being and that of society.

## Data Availability

The data underlying the results presented in the study are available are included in a separate file "Supporting Information"

## Acknowledgment

The authors would like to thank the DHS program for approving the use of these data to advance scientific knowledge on women’s autonomy in Tanzania.

